# Cohort Profile: Born in Wales - a birth cohort with maternity, parental, and child data linkage for life course research in Wales, UK

**DOI:** 10.1101/2023.06.14.23291389

**Authors:** HE Jones, MJ Seaborne, NL Kennedy, ML James, S Dredge, A Bandyopadhyay, A Battaglia, S Davies, S Brophy

## Abstract

**Purpose:** Parental and neonatal child health and education records have been linked to provide an entire country birth cohort, to examine what will improve the health and wellbeing of families growing up in Wales. Established in 2020, Born in Wales utilised data linkage techniques to connect information from the 2011 census with health, social care, and education routine data in the Secure Anonymised Information Linkage (SAIL) Databank. We present the descriptive data available in the linked database, emphasise the robust data security and governance frameworks, and present the future expansion plans for the database beyond its initial development stage.

**Participants:** Descriptive information from 2011 to 2023 has been gathered from SAIL. This comprehensive dataset comprises over 400,000 child electronic records. To augment this data, the Born in Wales and primary school surveys have contributed quantitative and qualitative responses.

**Findings to date:** The cohort comprises all children born in Wales since 2011, with follow-up conducted until they finish primary school at age 11. 2,500 parents and 30,000 primary school children have been recruited for enhanced data collection and linkage to the data spine. The child cohort is 51%: 49% female: male, and 6% are from ethnic minority backgrounds. When considering age distribution, 26.8% of children are under the age of 5, while 63.2% fall within the age range of 5-11.

**Future plans:** Born in Wales will expand by 30,000 new births annually in Wales, while including follow-up data of children and parents already in the database. Supplementary datasets complement the existing linkage, including primary care, hospital data, educational attainment and social care. Future research includes exploring the long-term implications of COVID-19 on child health and development, the influence of environmental factors including climate change on health and examining the impact of parental work environment on child health and development.

**Strengths and limitations of this study:**

- Born in Wales has established a comprehensive, Wales-wide population-based database which consolidates clinical data from maternity, neonatal, child health, and education records.
- This national-scale database is supplemented by quantitative and qualitative results from surveys conducted by Born in Wales, providing rich insights into details that cannot be obtained through routinely collected data.
- The existence of this database enables further data linkage, facilitating life course research on the health and wellbeing of the Wales population.
- Missing data or errors in routine and administrative data may be constraint.
- A potential restriction of Born in Wales is the loss of data pertaining to individuals who relocate outside of Wales during pregnancy or after the child’s birth.

## Introduction

The recognition of health promotion early in life is acknowledged as a strategy to enhance health and wellbeing across the life course [1]. This longitudinal approach encompasses the stages of pregnancy planning, pregnancy, childhood, adolescence, and adulthood [2]. Birth cohorts such as Born in Bradford (BiB) [3], the eLIXIR Partnership [4], and Born in Scotland [5] have contributed substantial evidence in the field of life course research, particularly focusing on pregnancy and early childhood.

Traditional cohorts are formed by recruiting participants within a specified timeframe. However, these cohorts can very quickly become outdated due to fluctuations in population demographics, lifestyle factors, and environmental variations. Follow-up of such cohorts becomes challenging, as sample attrition, where participants withdraw, leave, or do not complete the study, introduces methodological biases that can impact research validity and findings [6]. Furthermore, cohort recruitment may suffer from volunteer bias or self-selection bias, potentially limiting the representativeness of the cohort [7]. Population-based cohorts eliminate these biases and provide broader information about a wider scale of outcomes that may not be feasible in other cohort studies [8].

Previous birth cohorts have faced challenges related to attrition and inclusion bias. Routine electronic health records, which encompass data on all women receiving antenatal care in Wales and their infants, offer a solution to reduce these complications [9]. While national birth cohorts have been well established in other countries [10,11], the linkage of maternal and child data has not been extensively utilised in the UK. In Scotland, population data linkage has yielded findings that have informed UK clinical guidelines regarding maternal and neonatal outcomes [12]. Numerous successful linkages, such as maternity data with national birth registration datasets, birth registration with Hospital Episode Statistics (HES), and utilisation of primary care pregnancy data in the UK [13–17], have demonstrated the utility of this approach.

Born in Wales facilitates a platform for linking research datasets with authorised levels of patient anonymity and assured data security. Additional advantages include the capability to consolidate various datasets from other organisations, including healthcare, social care, and education. Ethical guidelines must be adhered to when working with these datasets to ensure accurate data linkage and participant anonymity.

Data-linkage facilitated by Born in Wales provides a unique data repository for investigating significant public health questions with far-reaching implications. The capacity to run these linkages permits a wide range of longitudinal health, social, and education data to be collected along with the ability to enhance life course data analysis. Born in Wales operates as a longitudinal database, commencing in pregnancy, and incorporating routinely collected clinical data from maternity, neonatal, social care, and education records, not solely relying on participant recruitment. Moreover, Born in Wales has the added element of supplementation by survey responses as well as the population-level data outcomes. With records of over 30,000 new births in Wales annually, Born in Wales has the potential to evolve into one of the most comprehensive datasets on parental and child health. The Secure Anonymised Information Linkage (SAIL) Databank [18,19] plays a vital role in integrating and harmonising these datasets.

This document outlines the safeguards implemented to protect user rights throughout the development and application of Born in Wales, as well as providing the demographics of the Born in Wales cohort. These procedures and mechanisms adopted are established on the successes observed in other databases and cohorts [3–5].

## Cohort description

### Data sources

Born in Wales establishes linkages between electronic data pertaining to mothers, babies, and partners (including self-identified biological fathers or mothers’ partners) for children born in Wales, encompassing approximately 30,000 annual births. The linked health records encompass primary care data (from Wales Longitudinal General Practice (WLGP)), secondary care (from hospital admissions, emergency care, inpatient from Patient Episode Database for Wales (PEDW), and outpatient from Outpatient Database for Wales (OPDW)), maternal indicators (midwife data), public health records (vaccination uptake, hearing checks, health visitor assessment, breastfeeding initiation and duration, and COVID-19 vaccination/testing) as well as a core database for the National Community Child Health Database (NCCH). Additionally, data pertaining to education, Census 2011, police/domestic violence, substance abuse, social care (looked after children, child protection register, children in receipt of care, and family court) have been successfully linked. Moreover, connections are being established to Census 2021 and the National Neonatal Audit Database. Ultrasound scans conducted across Wales have been coded to identify follow-up markers predictive of future cognitive development and school readiness, these have also been prepared for integration into the Born in Wales data spine. Maternity and neonatal data were acquired from the SAIL Databank, extracting from multiple datasets including the NCCH Dataset, Maternal Indicators Dataset (MIDS), and Patient Episode Dataset for Wales (PEDW). Please refer to Table 1 for further details regarding the datasets already linked or planned to be linked in Born in Wales.

In addition to the electronic birth cohort, the data spine is enhanced with repeated surveys administered to both mothers and identified biological fathers or female/male partners during pregnancy. These surveys capture self-reported health data pertaining to stress, mental health, occupation, ethnicity, and open ended questions addressing strategies to enhance health and wellbeing for families [20]. Over a 5 year period, an average of 1,000 women/families per year will be targeted for recruitment to further enrich the birth cohort. The enhanced data obtained from these surveys will be utilised to extrapolate (impute data) for the larger cohort study, enabling the estimation of variables such as income in the comprehensive all Wales cohort. It is important to note that 94% of the population in Wales is of Caucasian ethnicity.

**Table 1.**
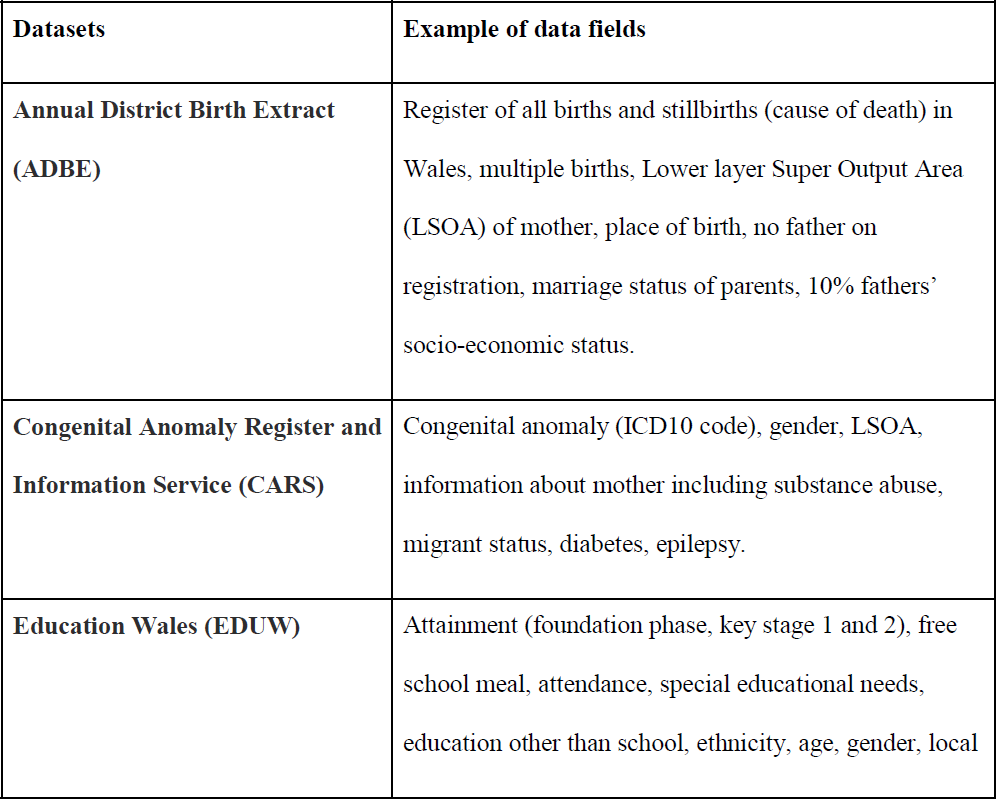
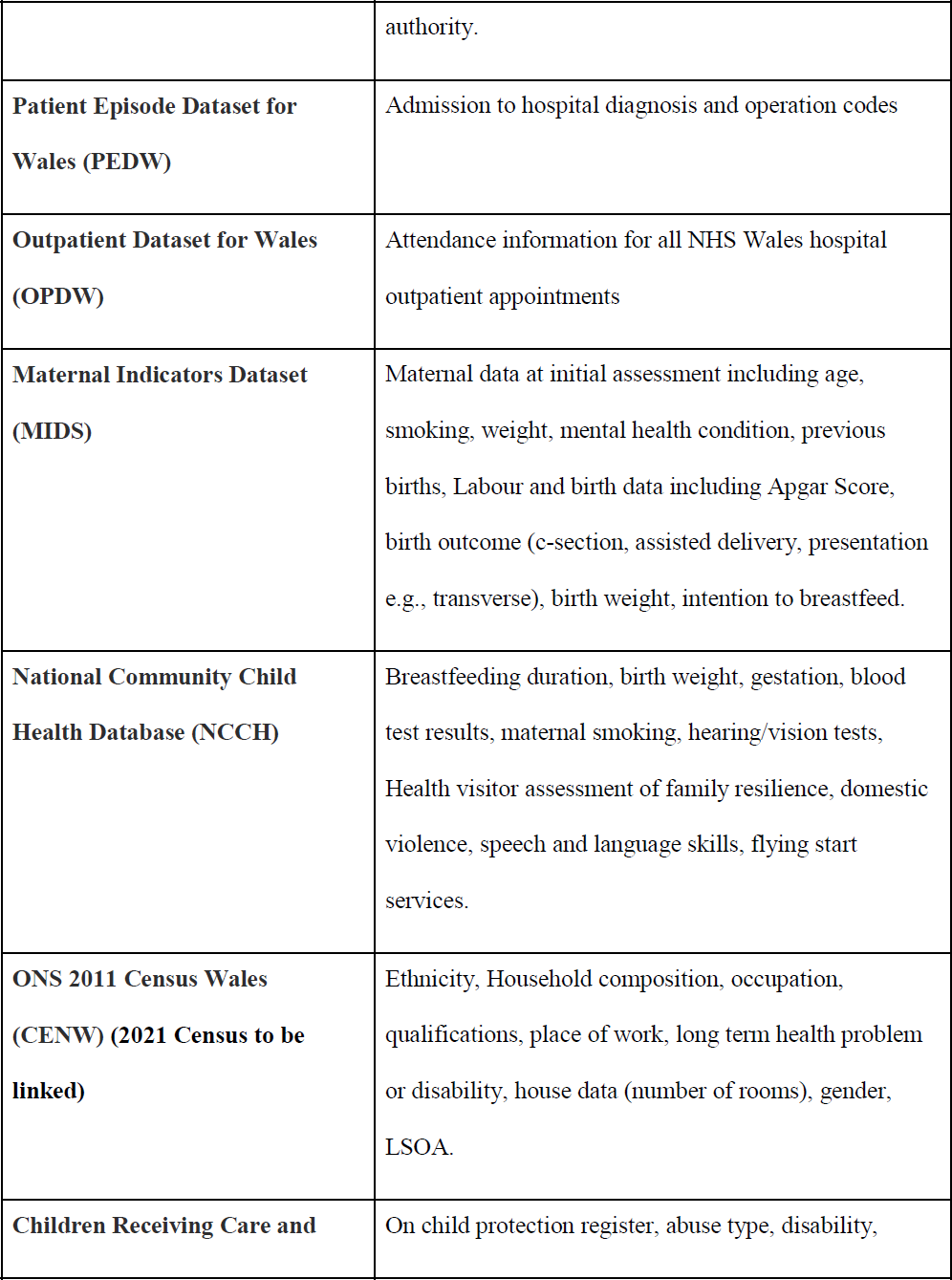
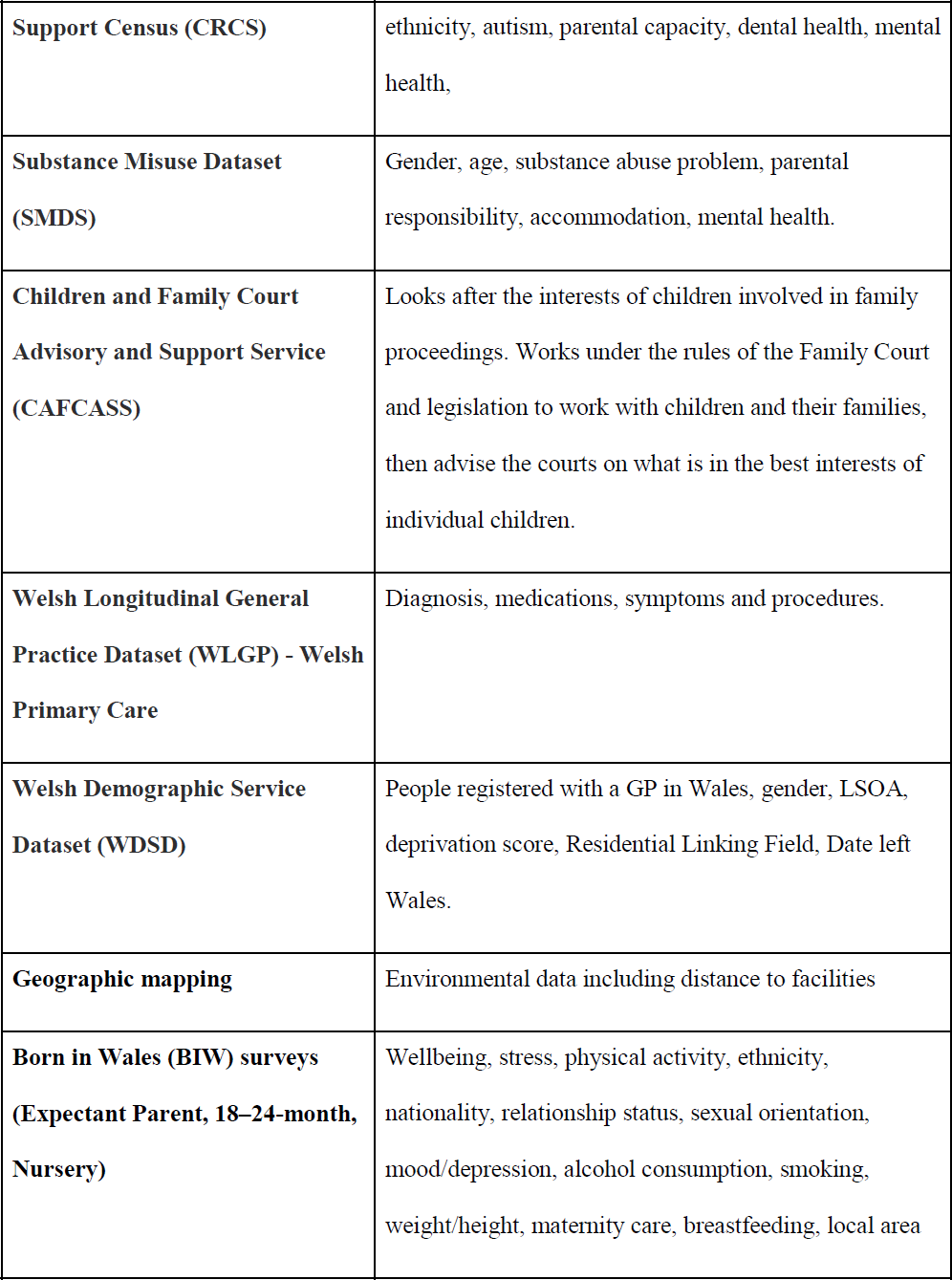
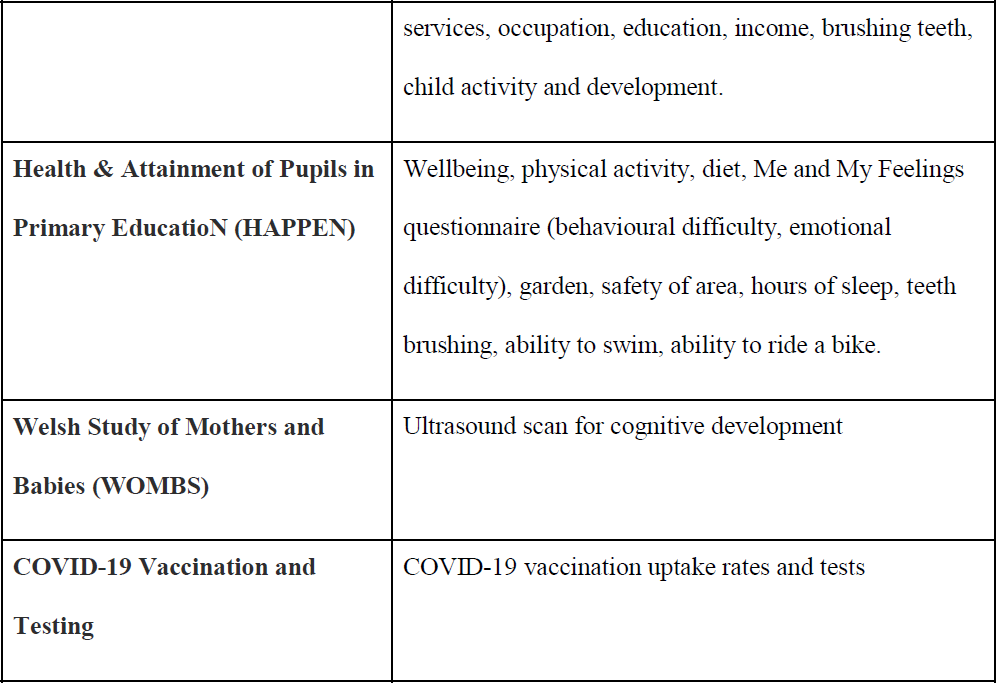
Datasets linked or to be linked in Born in Wales

### Data-linkage hosting environment

The core datasets of Born in Wales consist of PEDW, MIDS, and NCCH. These datasets are enhanced by linking them to HAPPEN, CENW, WOMBS, and Born in Wales survey data. Further linkages then can include multiple datasets including CARS, EDUW, CAFCASS and CRCS. The core components of Born in Wales and subsequent linkages are described in Figure 1. The foundation of Born in Wales involves SAIL’s robust anonymising system, which ensures secure data linkage in adherence to specified data protocols. As part of the data upload process to SAIL, the original dataset is divided into two distinct types of files. File 1 encompasses sensitive person-level demographics data, which is transmitted to Digital Health and Care Wales (DHCW). DHCW undertakes the processing, matching, and anonymisation of the File 1 data before transmitting it to SAIL. File 2 comprises clinical data or other non-identifiable data, which is directly transmitted to SAIL (See flow diagram of this process in Figure 2).

**Figure 1.**
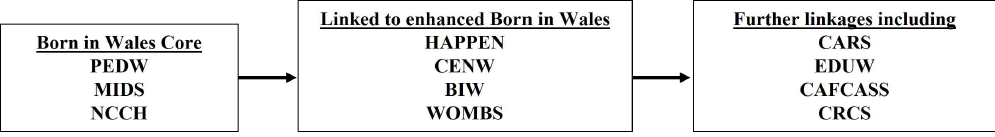
Core components of Born in Wales and further linkages

**Figure 2.**
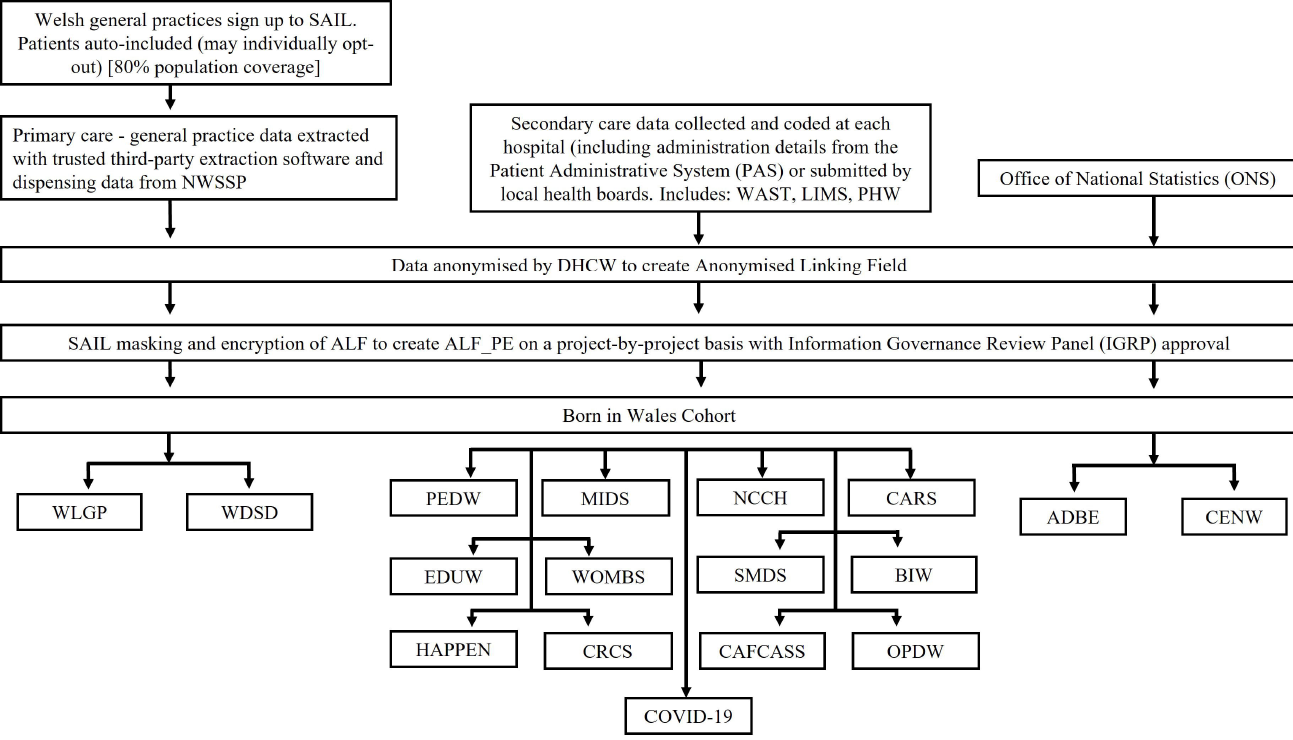
Flow diagram of the SAIL process

### Data-linkage procedures and resources

SAIL employs an Anonymous Linking Field (ALF) to establish connections with other records. Matching of two records arises when their respective ALFs are identical. Within the Born in Wales database, both maternal and child ALFs are utilised, enabling the identification and linkage of mothers to their corresponding children. Consequently, this integration culminates in the creation of a comprehensive data repository, accessible to researchers who can request extracted data encompassing maternity, neonatal, and/or mental health services. The matching algorithm was collaboratively designed and assessed by a trustworthy third party and SAIL. The algorithm compares numerous personal identifiers between the received dataset and the Welsh Demographic Service Dataset (WDSD). The linkage is performed based on an NHS number whenever possible (deterministic linkage). In the absence of an NHS number, a matching algorithm utilising surname, first name, post code, date of birth and sex is applied against the Welsh Demographic Service (WDS) Dataset (probabilistic linkage). The algorithm’s development ensures a high matching accuracy, with specific thresholds for match accuracy reported to SAIL within the anonymised dataset. A poor matching score indicates incomplete or inaccurate personal information provided by an individual or a lack of registration on the NHS database. The ALF system ensures a high level of match quality, as it facilitates the accurate matching of infants born within Born in Wales with their respective mothers’ records [18].

### Patient and public involvement

Patient and Public Involvement (PPI) is integral to the growth of Born in Wales, underscoring its commitment to inclusivity and participatory research. The inclusion of PPI ensures that members of the public actively participate in shaping the research project. Valuable input was sought during the design of the questionnaires, complemented by the insights provided by midwives who shared their lived experiences on pertinent research inquiries. Moreover, a dedicated PPI steering group will be actively sought to incorporate feedback from the public. The PPI group will be recruited from midwives and health visitors via already established networks, along with expectant and new parents to help co-produce its delivery and approach from the survey to research questions and aims. Born in Wales aims to adopt a co-production approach, emphasising collaborative partnerships among researchers, practitioners, and the public. The decision-making processes within Born in Wales will prioritise PPI to ensure the inclusion of diverse perspectives. To facilitate this approach, Born in Wales will follow the Co-production of Research and Strategy (CORDS) standard operating procedure [21] and the UK standards for PPI involvement and guidance from INVOLVE [22] which establishes a framework for meaningful engagement with practitioners and families. This approach fosters a reciprocal exchange, focusing on the important questions and concerns for stakeholders, while also enhancing the impact, translation, and dissemination of findings. A record of PPI activity will be maintained using the Public Involvement in Research Impact Toolkit (PIRIT) [23].

## Findings to date

### Born in Wales cohort characteristics

The cohort comprises all children born in Wales since 2011 and followed until age 11. An electronic data spine has been established to encompass the entire population of children born in Wales in the last 13 years, resulting in a cohort size exceeding 400,000 children within Born in Wales. Furthermore, 2,500 parents and 30,000 primary school children have been recruited to facilitate enhanced data collection, all of whom are linked to the electronic cohort data spine; ongoing recruitment efforts are underway. This cohort encompasses the entirety of Wales, thus represents a national birth cohort for the country. In terms of gender distribution, females comprise 51% of the cohort, while males account for 49%, additionally, 6% of the cohort are from an ethnic minority background. Analysis of the NCCH Births dataset reveals that 26.8% of children are below the age of 5, whereas 63.2% are aged between 5-11 years. Between 2011 to 2022, a total of N=435,032 births were recorded, involving N=269,427 mothers. Of these births, N=222,867 are male, N=212,188 are female births, and N=40 the gender data is missing.

### Maternal, birth and birth outcomes

Between 2011 to 2022, there were N=1,718 stillbirths, constituting 0.4% of all births. Examining gestational age, 7.26% (N=31,625) of infants were born preterm, defined as delivery before 37 weeks gestation, while 82.8% (N=360,825) were born at term or later (37 weeks and onwards). The Born in Wales database has recently been utilised to investigate the risk factors associated with low birth weight (LBW). Analyses revealed that non-singleton children exhibited the highest risk of LBW (OR 21.74 (95% CI 21.09 to 22.40)), followed by pregnancy intervals of less than 1 year (OR 2.92 (95% CI 2.70 to 3.15)). Maternal physical and mental health conditions, including diabetes (OR 2.03 (95% CI 1.81 to 2.28)), anaemia (OR 1.26 (95% CI 1.16 to 1.36)), depression (OR 1.58 (95% CI 1.43 to 1.75)), serious mental illness (OR 1.46 (95% CI 1.04 to 2.05)), anxiety (OR 1.22 (95% CI 1.08 to 1.38)) and use of antidepressant medication during pregnancy (OR 1.92 (95% CI 1.20 to 3.07)) were also identified as significant risk factors for LBW. Additional maternal risk factors include smoking (OR 1.80 (95% CI 1.76 to 1.84)), alcohol-related hospital admission (OR 1.60 (95% CI 1.30 to 1.97)), substance misuse (OR 1.35 (95% CI 1.29 to 1.41)) and evidence of domestic abuse (OR 1.98 (95% CI 1.39 to 2.81)) [24].

### Born in Wales survey participants

Among the participants of the Born in Wales survey, the mean age was 32 years (SD=12.5), with an interquartile range (IQR) of 28-35 years. The majority of survey participants identified as white ethnicity, accounting for 45.5% of the sample (51% did not disclose their ethnicity). The Born in Wales survey data was utilised in a mixed methods study conducted during the COVID- 19 pandemic, aiming to examine the effects of the pandemic on pregnancy experiences and birth outcomes in 2020. Results indicated that the pandemic had a notable adverse effect on the psychological wellbeing of 71% of survey respondents. These individuals reported heightened levels of anxiety, stress, and feelings of loneliness, which were associated with attending prenatal scans in the absence of their partner, giving birth alone, and restricted interactions with midwives. However, no significant differences were observed in annual birth outcomes, including gestation, birth weight, stillbirths, and Caesarean sections, between infants born in 2020 compared to 2016–2019. Whilst the pandemic negatively affected mothers’ pregnancy experiences, population-level data indicates that this did not result in adverse birth outcomes for infants born during the pandemic [25].

### HAPPEN children’s primary school network

Health & Attainment of Pupils in Primary EducatioN (HAPPEN) represents a substantial cohort of over 30,000 school-aged children across Wales, with a gender distribution of approximately 47% boys, 49% girls and 3% who preferred not to disclose their gender. The average age of the children is 9.35 years. Recently, the HAPPEN dataset has been utilised to explore the health and wellbeing of children during the COVID-19 pandemic. This encompassed a retrospective cohort study employing an online cohort survey conducted between January 2018 to February 2020, in conjunction with routine PCR SARS-CoV-2 test results. The study explored health-related behaviours in children spanning the period from 2018 to 2020, and their association with being tested and testing positive in 2020 to 2021. Notably, the investigation revealed significant associations between parental health literacy and monitoring behaviours [26]. Furthermore, the study employed free school meal status (FSM) as a proxy for deprivation in an exploratory analysis examining the impact of school closures on the health and wellbeing. The findings indicated that children eligible for FSM may experience adverse consequences in terms of physical health, including reduced physical activity and suboptimal dietary choices, as a result of prolonged school closures [27].

## Discussion

### Strengths and limitations

Born in Wales has established a comprehensive, Wales-wide population-based database which consolidates clinical data from maternity, neonatal, child health, and education records. This national-scale database is supplemented by quantitative and qualitative results from surveys conducted by Born in Wales, providing rich insights into details that cannot be obtained through routinely-collected data. The existence of this database enables further data linkage, facilitating life course research on the health and wellbeing of the Wales population. This has significant implications for enhancing healthcare delivery at the local level and offering valuable research evidence to guide policy and practice both within the studied population and in other countries. This research aims to generate robust evidence that can influence policy and practice, enabling the implementation of early interventions to promote child health and wellbeing. Moreover, collaborations with diverse cohorts facilitate comparisons across different populations, enabling a broader understanding of factors influencing child health and wellbeing outcomes.

One inherent constraint associated with routine and administrative data is the presence of missing data or errors. However, SAIL has established a robust framework that includes a team of skilled analysts and rigorous quality control procedures to address issues such as duplication of patient data entries and minimise instances of missing data. Subsequently, the Born in Wales project can contribute to the enhancement of clinical reporting practice and bolster the reliability of research findings derived from the database. Additionally, the utilisation of anonymised cohorts serves as an effective strategy for overcoming the barriers related to obtaining consent from individuals, thereby facilitating the seamless aggregation and analysis of data.

A potential constraint of Born in Wales is the loss of data pertaining to individuals who relocate outside of Wales during pregnancy or after the child’s birth. Analogous to other datasets, research conducted utilising Born in Wales may be constrained by the information available through routine data collection. Nevertheless, Born in Wales can enhance the available information by incorporating survey questions that capture data missing from administrative records. It is important to acknowledge that data entry can be subject to human error, and there may still be instances of missing records.

### Research aspirations

As the Born in Wales database continues to expand, there are intentions to broaden the scope by incorporating additional health, social care, and education data, alongside Police data. The subsequent phase of data linkage will involve integrating Police data into SAIL. Replicating this data linkage model on a national scale within the UK and potentially extending it globally would enable the establishment of more extensive research cohorts and facilitate cross-comparisons across diverse populations.

This cohort is established through the integration of routine data records, encompassing health and administrative data, as well as environmental data supplemented with survey data to enrich the cohort. It constitutes a comprehensive, data rich total population cohort constructed through substantial investments and extensive record linkage efforts from HDR UK and ADR. Furthermore, this methodology demonstrates cost-effectiveness by leveraging existing data sources, establishing a dynamic cohort that continuously expands its participant pool. The cohort is well suited for investigating natural experiments and interventions. The endeavours undertaken in this study closely align with recommendations 1-3 and 6 of the MRC strategy to maximise UK population cohorts [28].

The design of this cohort facilitates cross-cohort comparisons with other electronic cohorts in the UK, including Born in Bradford [3] and eLIXIR [4]. To harmonise phenotypic variables across studies and adopt core common data standards, all data in the cohort will adhere to the OMOP standard. Knowledge sharing and directories have been facilitated through the HDR UK gateway, and high-quality, robust meta-data is provided, adhering to recommendations 4 and 5 of the MRC strategy [28].

This cohort is unique due to the comprehensive breadth of routine and survey data collected and linked in a total population cohort. It is designed to enable data sharing by employing harmonised data and meta-data, following the principles of findability, accessibility, interoperability, and reusability (FAIR). A distinguishing feature of Born in Wales is its incorporation of census data linked to numerous datasets, encompassing social care and justice datasets. Additionally, the database includes self-reported data which captures aspects that are not typically electronically captured, such as stress, wellbeing, physical activity, access to services, and use of services.

### Data availability and access

The data for the Born in Wales cohort is available in the SAIL databank at Swansea University, Swansea, UK [29]. All proposals to use SAIL data are subject to review by an independent Information Governance Review Panel (IGRP). This project’s approval code is 0916. Before any data can be accessed, approval must be given by the IGRP. To use Born in Wales data you need to provide a safe researcher training certificate, a signed data access agreement and IGRP approval.

### Future directions

A comprehensive research database has been established, encompassing data pertaining to maternity, neonatal, and child health. This database integrates information on maternal and paternal health throughout pregnancy and extends to encompass the subsequent health of the child. Moreover, it enriches the dataset with valuable qualitative insights obtained from the Born in Wales surveys, which capture nuanced information not captured solely by hospital records. Born in Wales is committed to continuous progress and expansion through collaborative efforts, enabling longitudinal follow-up of families in Wales and fostering co-production of research questions relevant to each stage of development. By encompassing the entire population of Wales, Born in Wales generates a wealth of rich data on the national scale, which has the potential to inform interventions aimed at promoting healthy growth and development among the population.

## Data availability statement

Data are available upon reasonable request. Researchers can apply for data access by submitting a research application to the SAIL team. The SAIL website provides information on the application process (https://saildatabank.com/data/apply-to-work-with-the-data/). All proposals to use SAIL data are subject to review by an independent Information Governance Review Panel (IGRP). This project’s approval code is 0916. Before any data can be accessed, approval must be given by the IGRP. To use Born in Wales data you need to provide a safe researcher training certificate, a signed data access agreement and IGRP approval.

## Contributors

HJ, SB and SD contributed to the conception of this article. MS, NK, MJ, AB, AB, and SD were involved in manuscript writing and revision. HJ, MS, AB, and MJ were involved in data analysis and interpretation. All authors read and approved the final manuscript.

## Patient and public involvement

Patients and/or the public were involved in the design and dissemination of this research and a PPI steering group is being sought.

## Data Availability

Data are available upon reasonable request. Researchers can apply for data access by submitting a research application to the SAIL team. The SAIL website provides information on the application process. All proposals to use SAIL data are subject to review by an independent Information Governance Review Panel (IGRP). This projects approval code is 0916. Before any data can be accessed, approval must be given by the IGRP. To use Born in Wales data you need to provide a safe researcher training certificate, a signed data access agreement and IGRP approval.

https://saildatabank.com/data/apply-to-work-with-the-data/

